# Aerosol and bioaerosol particle size and dynamics from defective sanitary plumbing systems

**DOI:** 10.1101/2020.11.01.20223974

**Authors:** Michael Gormley, Thomas J Aspray, David A Kelly

## Abstract

Aerosols are readily transported on airstreams through building sanitary plumbing and sewer systems and those containing microbial pathogens (known as bioaerosols) are recognised as contributors to infection spread within buildings. When a defect occurs in the sanitary plumbing system that affects the system integrity, a cross-transmission route is created that can enable the emission of bioaerosols from the system into the building. These emission occurrences are characterised as short-burst events (typically < 1 minute in duration) which makes them difficult to detect and predict. The characterisation of these emission events is the focus of this research.

Two methods were used to characterise bioaerosol emission events in a full scale test rig: (i) an Aerodynamic Particle Sizer (APS) for particle size distribution and concentrations; and (ii) a slit-to-agar sampler to enumerate the ingress of a viable tracer microorganism (*Pseudomonas putida)*. The APS data confirmed that most particles (> 99.5%) were <5 μm and were therefore considered aerosols. Particles generated within the sanitary plumbing system as a result of a toilet flush leads to emissions into the building during system defect conditions with an equivalence of someone talking loudly for over 6 and a half minutes. There were no particles detected of a size > 11μm anywhere in the system. Particle count was influenced by flush volume, but it was not possible to determine if there was any direct influence from airflow rate. Typical emissions resulting from a 6 litre flush were in the range of 280 – 400 particles per second at a concentration of typically 9 to 12 number per cm^3^ and a total particle count in the region of 3,000 to 4,000 particles, whereas the peak emissions from a 1.2 litre flush was 60 - 80 particles per second at a concentration of 2.4 to 3 number per cm^3^ and a total particle count in the region of 886 to 1045 particles. The reduction in particles is in direct proportion to the reduction in flush volume. The slit-to-agar sampler was able to provide viable time course CFU data and confirmed the origin of the particles to be the tracer microorganism flushed into the system. The time course data also has characteristics consistent with the unsteady nature of a toilet flush.

## 1 INTRODUCTION

### 1.1 Background

The mechanism of airborne transmission of the SARS-CoV-2 virus, specifically the transport of virus-laden particles (bioaerosols) in the air, has been a source of much debate since the virus was first identified in Wuhan, China, in December 2019 ^1^. The rapid spread of the SARS-CoV-2 virus around the world has caused a global coronavirus disease (COVID-19) pandemic.

Most of the focus on how the SARS-CoV-2 virus is transmitted has concentrated on human-to-human transmission. This is largely due to a general lack of knowledge of how infections spread ^2^. Generally, viral respiratory infections are considered to be spread by direct contact, by touching an infected person or the surfaces and other fomites that the infected person has touched ^3^. Bioaerosols expelled by the infected person can land on surfaces where the virus can remain stable for days ^4^. Droplets can also be deposited directly onto a person in close proximity to the infected person. It is on this basis that frequent hand-washing and maintaining social distance are considered the main precautions against contracting infection. The transmission route which is less well understood, and often overlooked, is the transport of the virus in the air as bioaerosols, which can be so small that they can travel long distances within airstreams.

Generally, droplets are referred to as particles with a diameter >5 μm that fall rapidly to the ground under gravity, and so are transmitted only over a limited distance (e.g. ≤1 m), see Table 1. Aerosols, on the other hand, are particles with a diameter ≤5 μm that can remain suspended in the air for significant periods of time, allowing them to be transmitted over longer distances >1 m ^5–7^ Bioaerosols is the term used to define aerosols that contain viruses, bacteria, fungi, or bacteria and fungi cell components ^8^. They can also contain pollen grains or other biological material.

**Table 1:**
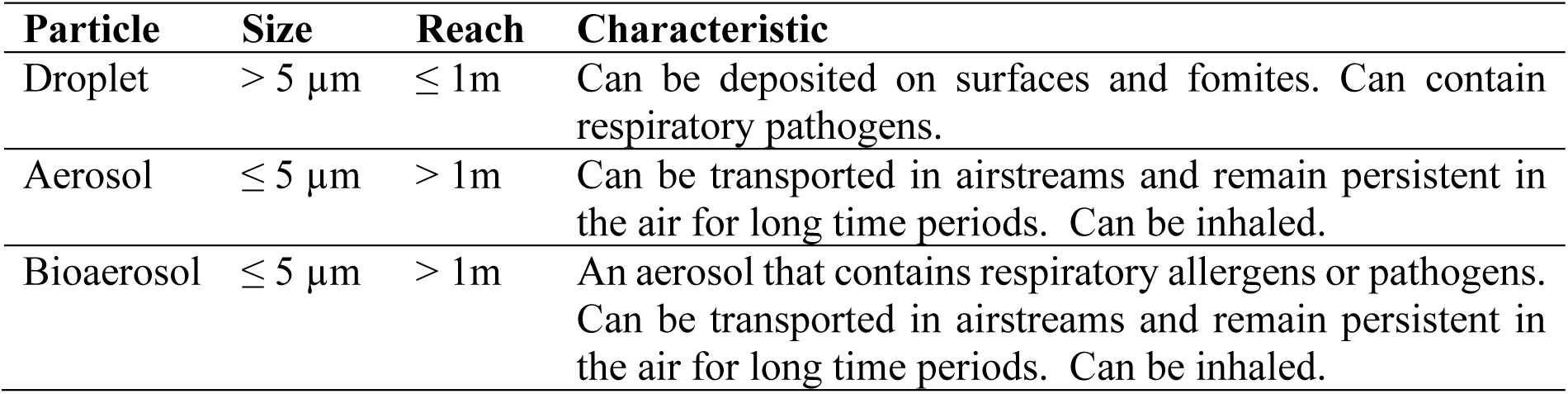
Definitions and characteristics of particles of different sizes.

The research presented in this paper investigates the aerosolisation of pathogens inside the sanitary plumbing system and their transmission as bioaerosols into the building during system defect conditions. Literature on this topic is very sparse since most research to date has focused on the ‘room side’ rather than the ‘sanitary plumbing side’. Work on flushing toilets Verani *et al*., Best *et al*., Johnson *et al*., Noble *et al*., Gebra *et al*., and Baker *et al*. ^9–11^ all present compelling evidence on the role that a toilet flush can play in generating bioaerosols within the bathroom. Our research differs from those cited in that it focuses on the aerosolisation process that occurs *inside* the sanitary plumbing system following wastewater discharge into the system, and the subsequent *emission* of aerosols (specifically bioaerosols) into the building through defects that can occur within the U-bend trap seal due to evaporation or the propagation of high pressure air surges inside the sanitary plumbing system ^12^. This is particularly relevant in tall buildings where sanitary plumbing system airflows and pressures can be relatively very high^13–16^.

The mechanism of aerosolisation within a sanitary plumbing system is due, in the main, to the unsteady discharge of wastewater into the system under normal usage. The surge waves generated as a result of these discharges play a large part in the aerosolisation process. One of the main surge generators within a sanitary plumbing system is the toilet. A 6 litre surge wave is typically generated with every flush and this interacts with the airflow and air pressures within the system to instigate aerosolisation. The characteristics of toilet flushes are well understood ^17^. Figure 1 illustrates the mechanism of aerosol generation within the sanitary plumbing system during and following toilet flushing.

**Figure 1:**
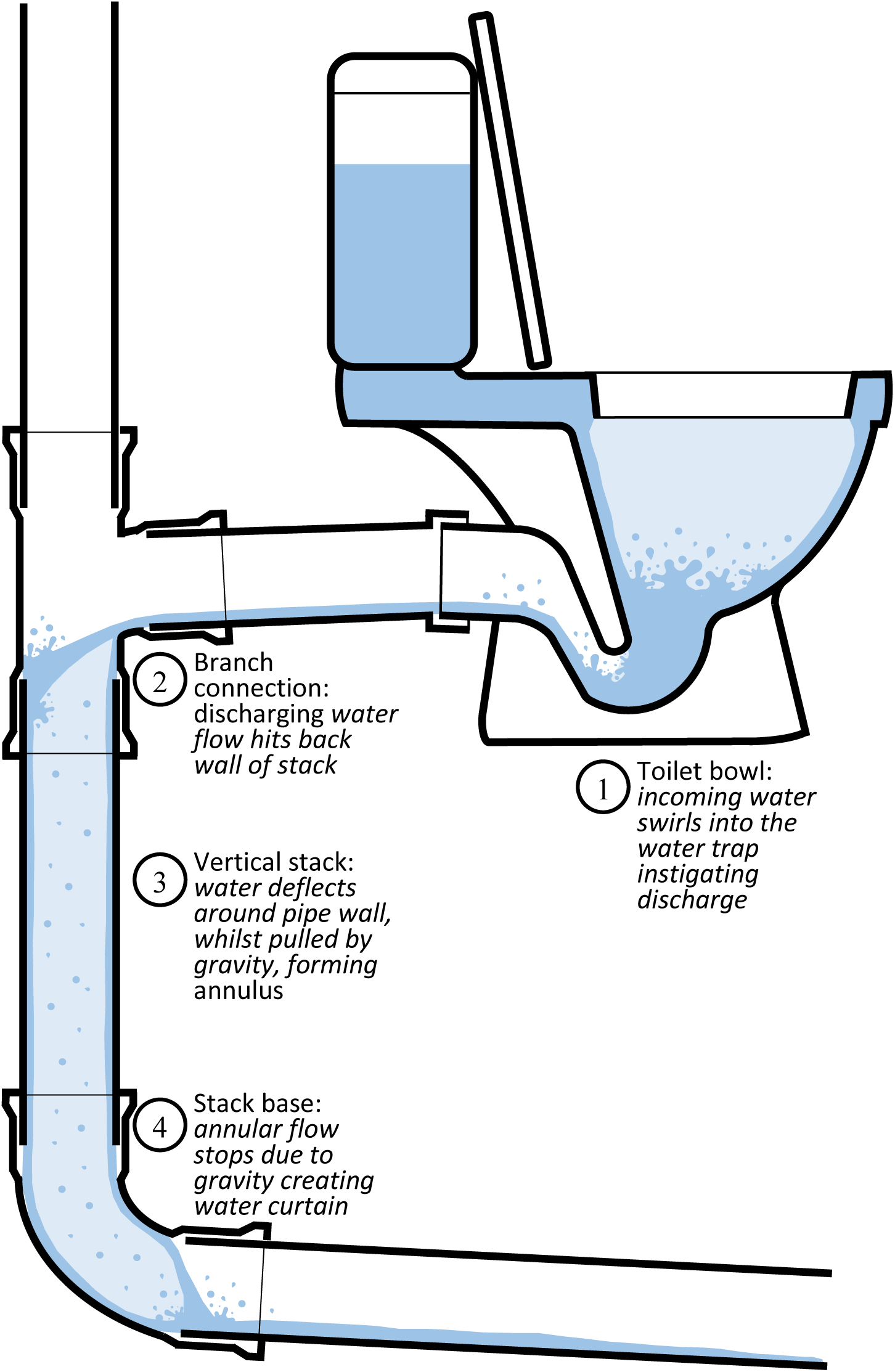
Mechanisms of aerosol generation during toilet flushing.

Whilst greater evidence surrounding the prevalence and infectivity of SARS-CoV-2 in faeces and wastewater is needed, the evidence is building and some studies have suggested that faeces can contain viable virions ^18,19^.

Recent work by Kang *et al*. ^20^ reports a probable case of local transmission of SARS-CoV-2 via defects within a building’s sanitary plumbing system. The sanitary plumbing system has been shown to be both a reservoir for pathogens ^21^ and a cross-transmission route between different areas within a building ^22^.

The ingress of bioaerosols into the building occurs following a pulsed air flow event. Such pulses are instigated by the dynamic response of airflows within the sanitary plumbing system to random sanitary appliance discharges. In experiments carried out by Gormley *et al*. (6), a two storey sanitary plumbing system was built and the extent of contamination was established. The pipe network, the airflows, the room surfaces, the room extract fan and associated ductwork were all found to be contaminated due to a single toilet flush. The flush contained a culture of *Pseudomonas putida* acting as a tracer organism.

Understanding these concepts has helped identify the source of specific outbreaks, notably the SARS-CoV-1 outbreak in one particular apartment block in Hong Kong in 2003 ^23–26^ where the sanitary plumbing system was implicated in the spread of the virus.

Figure 2 provides a graphical representation of the main modes of disease transmission, including the accepted human-to-human transmission under close contact, as well as the faecal-oral transmission modes via the mechanisms of room-side toilet flushing, and the subsequent system-side bioaerosol generation and ingress through a defective open trap. As studies have aimed to quantify and characterise the aerosol particle release from people when breathing, talking, singing, coughing, etc., this research aims to quantify and characterise the aerosol particle emissions from within a sanitary plumbing system following a toilet flush event.

**Figure 2:**
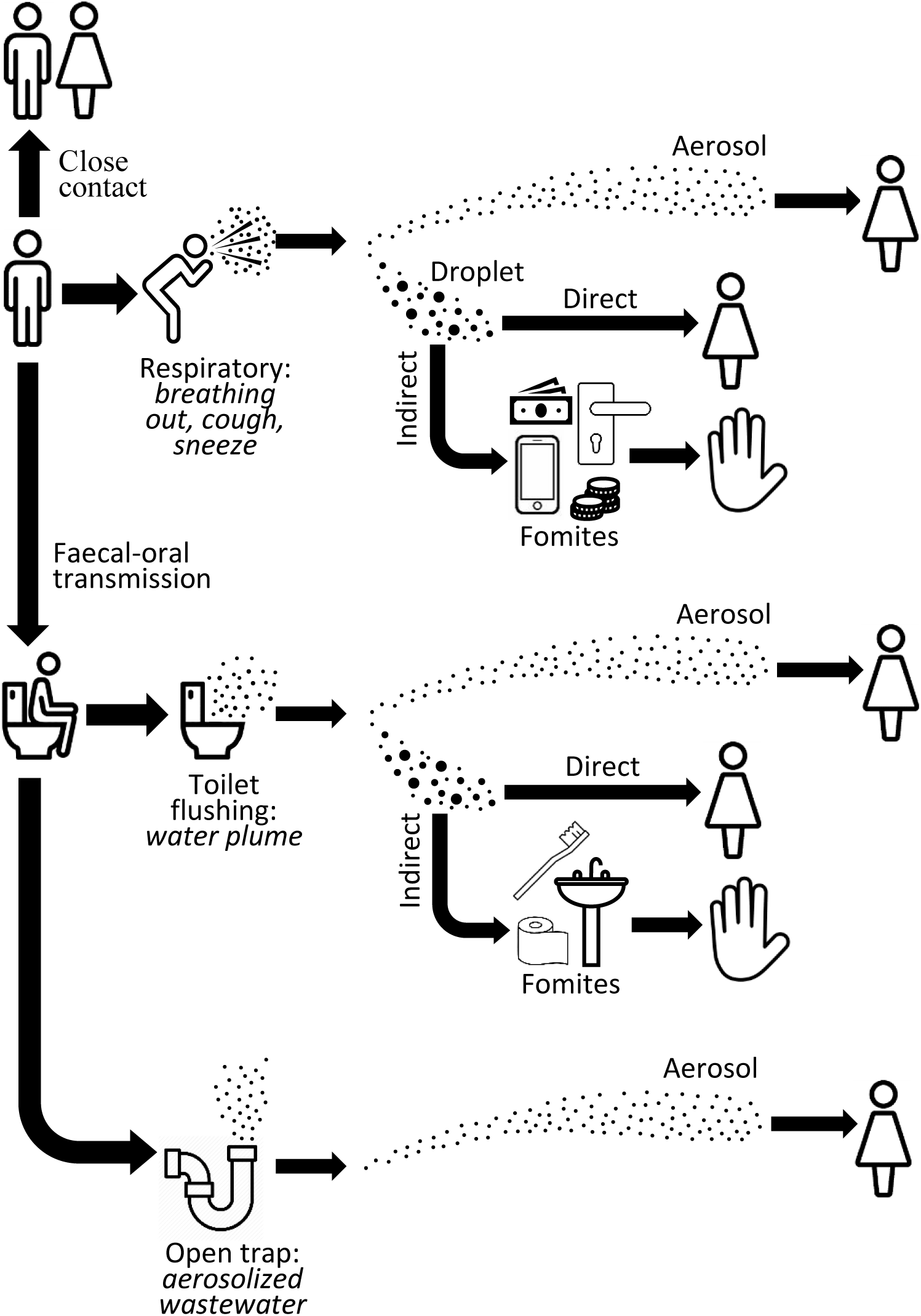
Aerosol generating mechanisms/processes and disease transmission routes within the built environment.

### 1.2 Paper Structure

Particle sizes, concentrations, and distributions are presented from the Aerodynamic Particle Sizer (APS) data. This represents what can be emitted into the building if there is a defect in the sanitary plumbing system. The intention is not to enumerate microorganisms in the air but to suggest the aerodynamic size of particles which leads to an understanding of what their impact might be on the transmission of contaminated air from the sanitary plumbing system into the building. The second dataset reported shows the timing and dynamics of aerosols emitted into the building via a defective U-bend trap seal having been generated following a simulated toilet flush which had been contaminated with a tracer organism, *P. putida*.

## 2 MATERIALS/METHODS

### 2.1 Test Rig

The test rig shown in Figure 3 was constructed in the Water Laboratory at Heriot-Watt University. The test rig is designed to European standards using EN12056:2000-2 ^27^ and represents a two storey building sanitary plumbing system. The system simulates a toilet flush on Floor 1 and has a sealed test chamber on Floor 2 (to represent a bathroom). The test chamber has an extract fan with a length of flexible ducting attached. Instruments for analysis of aerosols and bioaerosols (see below) through the system were positioned at the end of the ducting.

**Figure 3:**
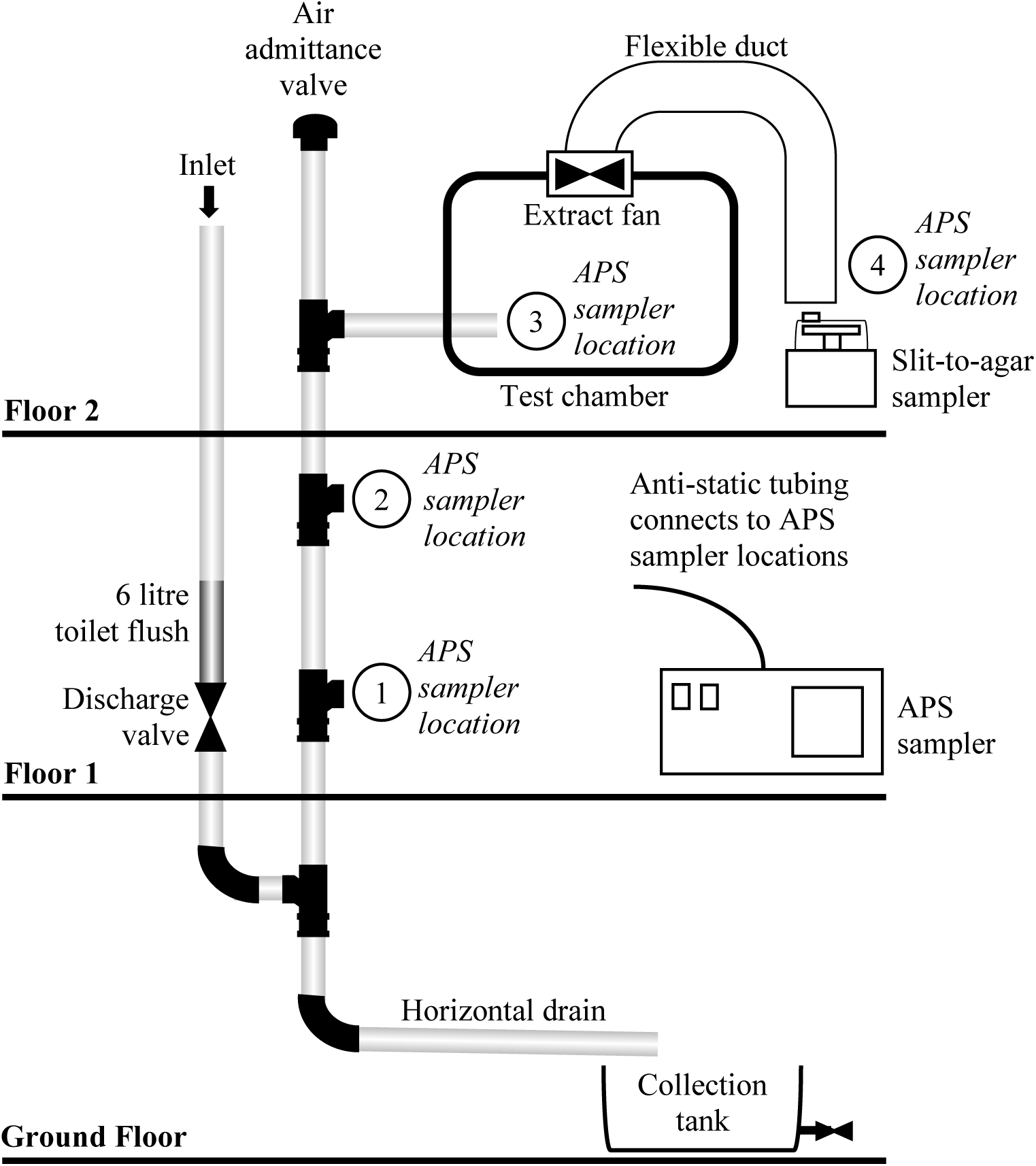
Building drainage system test-rig designed to BS EN12056:2000-2^27^.

The induced airflow in the system was set by a variable fan speed controller and the air velocity was measured by the anemometer in the pipework. To simulate a toilet flush, a pipe with a capacity to hold 6 litres of water (the maximum flush volume of a new toilet) was connected above a manual discharge valve. A 6 litre culture of *P. putida*, was inoculated into the drainage system via toilet flush and tracked at the outlet duct using the slit-to-agar sampler. The simulated toilet flush was initiated by opening the discharge valve, replicating the process in a normal drop valve flush toilet.

The discharge flow profile of a 6 litre drop valve toilet is shown in Figure 4. The flush event can be seen to last no more than 10 seconds. The fluid from the flush enters the vertical stack of the sanitary plumbing system and falls creating an annular flow. The shear between the water and the air causes an airflow to be entrained, thus causing air pressure fluctuations as the system attempts to balance airflow and pressure. The dynamics of this air intake follow the same pattern as the toilet discharge flow profile. The turbulence caused by the air/water mixture as they interact is sufficient to produce aerosols within the sanitary plumbing system. If the discharging wastewater contains pathogenic micro-organisms, then bioaerosols can be generated that can be transported on internal airflows.

**Figure 4:**
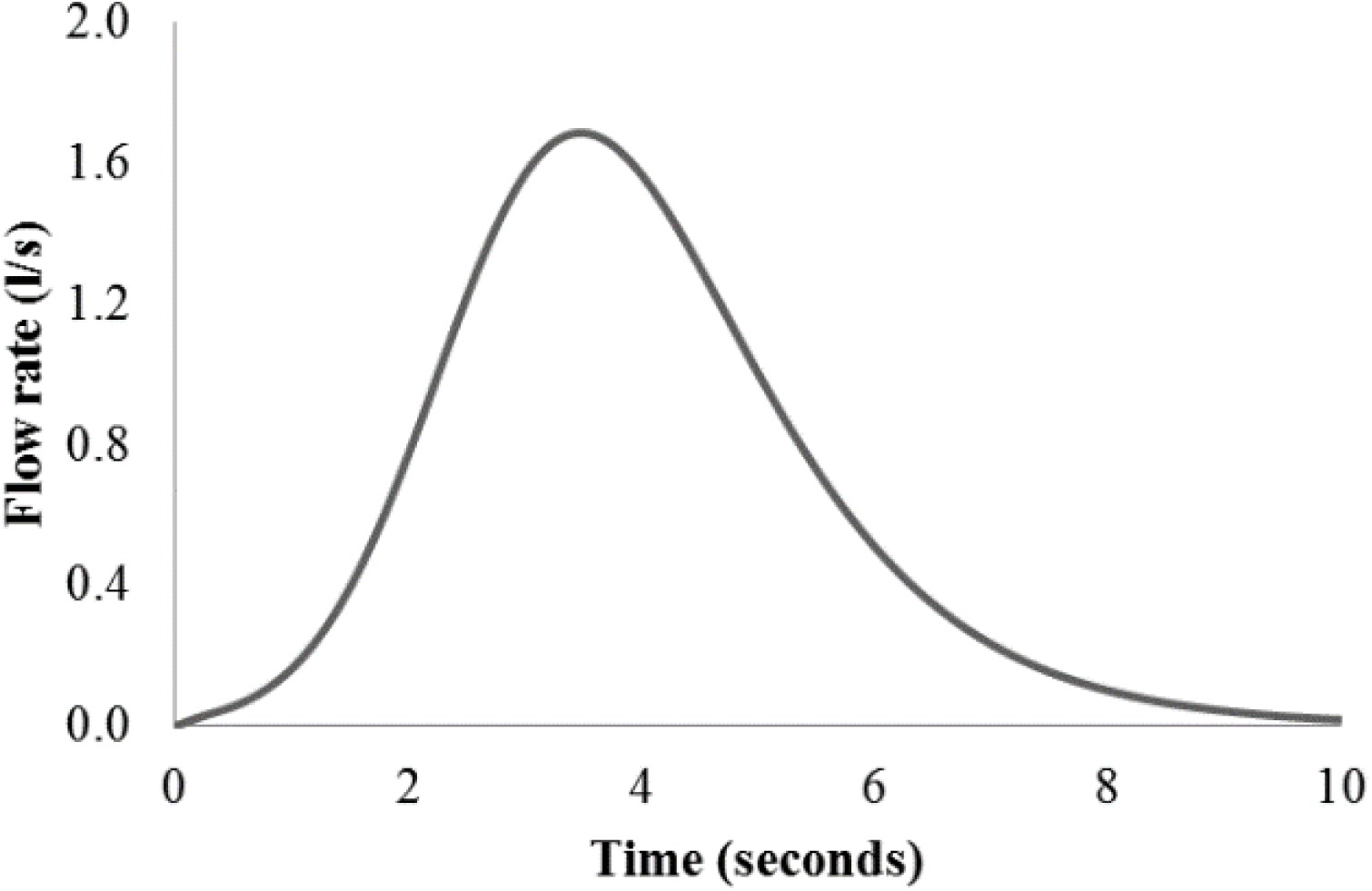
Discharge flow profile of a typical drop valve toilet.

### 2.2 Airflow

Since the test rig was in a laboratory (and not connected to a complete sanitary plumbing system or main sewer network) it was necessary to induce an updraught airflow using an extractor fan in the test chamber. The airflow rates used were defined previously ^22^ and were set to three broad ranges:

- Low flow rate (∼3 l/s)
- Medium flow rate (∼5 l/s)
- High flow rate (>6 l/s)

This ensured that results could be compared if they fell within the same range and ensured that flow rates were representative of those found in real systems.

### 2.3 Tracer organism

The class 1, non-genetically modified bacterium *P. putida* KT2440 ^28^ was used as a tracer organism. Pure colonies were picked from agar plates, inoculated into Erlenmeyer flasks containing tryptone soya broth and incubated overnight at 30 °C with an orbital shaking. The resulting stock culture was diluted to achieve an optical density at 600 nm wavelength (OD600) of 1.0 in 0.85 % (w/v) NaCl to provide an inoculum for the sanitary plumbing system (corresponding to ∼10^6^ cfu ml^-1^) forming the toilet flush volume.

### 2.4 Aerosol and bioaerosol monitoring

Two methods were used to sample and analyse the air particles emitted from the defective sanitary plumbing system: *i) Aerodynamic Particle Sizing (APS); and ii) Slit-to-agar air sampling*

#### i) Aerodynamic particle sizing (APS)

An APS device (model 3321, TSI Inc., Shoreview, MN) was used to enumerate the particle sizes of the air emitted from the test rig. The APS device was set to sample for 60 seconds before the airflow was initiated and the toilet flush was discharged. Samples were taken from the main vertical stack (APS sampler locations 1, 2, and 3 in Figure 3) and at the ductwork exit (APS sampler location 4 in Figure 3). Each test was set to record data for approximated 500 seconds. The data was recorded and processed to give time stamped recordings of specific particle sizes and the results graphed for inspection and analysis.

#### ii) Slit-to-agar sampler

A slit-to-agar air sampler (220 model; Mattson-Garvin) was setup at the end of the ductwork exit to monitor viable bioaerosols. The sampler was installed with the manufacturers’ 5 minute motor and set to sample at a flow rate of 65 standard cubic feet per minute (SCFM).

Before each experiment, the sampler plate housing and slit inlet were sterilised with 95% ethanol. A single 150 mm diameter Pseudomonas Isolation Agar (PIA) plate was placed on the rotor platform and secured with adhesive tape to prevent slippage. The height from plate to the slit was adjusted as required to allow for differences in agar thickness.

The sampler was set to run for 60 seconds before the toilet flush was discharged. The sampler was then allowed to run for a total of 300 secs per experiment. After the experiments, the PIA plates were incubated for 24 h at 30 °C before imaging and image analysis.

### 2.5 Image analysis

Images (.tiff format) of the slit-to-agar plates were opened in the image processing software ImageJ (http://imagej.nih.gov/ij). The angle tool was used to determine the time location of each individual colony.

### 2.6 Experimental procedure

The experimental procedure for both of these methods can be summarised as follows:

1. The system was washed down with biochlor^®^ solution and rinsed with tap water
2. The ‘inoculum’ was loaded into the valve controlled discharge pipe
3. The induced airflow was initiated and allowed to stabilise
4. The sampler (APS or slit-to-agar) was turned on and allowed to run for at least 60 seconds
5. The flush valve was opened to allow the ‘inoculum’ to discharge into the vertical stack Results were recorded in the APS, and for the slit-to-agar sampler, the PIA plate was marked up and taken for incubation

A range of ‘inocula’ were used to establish the importance of different factors. Table 2 shows the range of tests carried out.

**Table 2:**
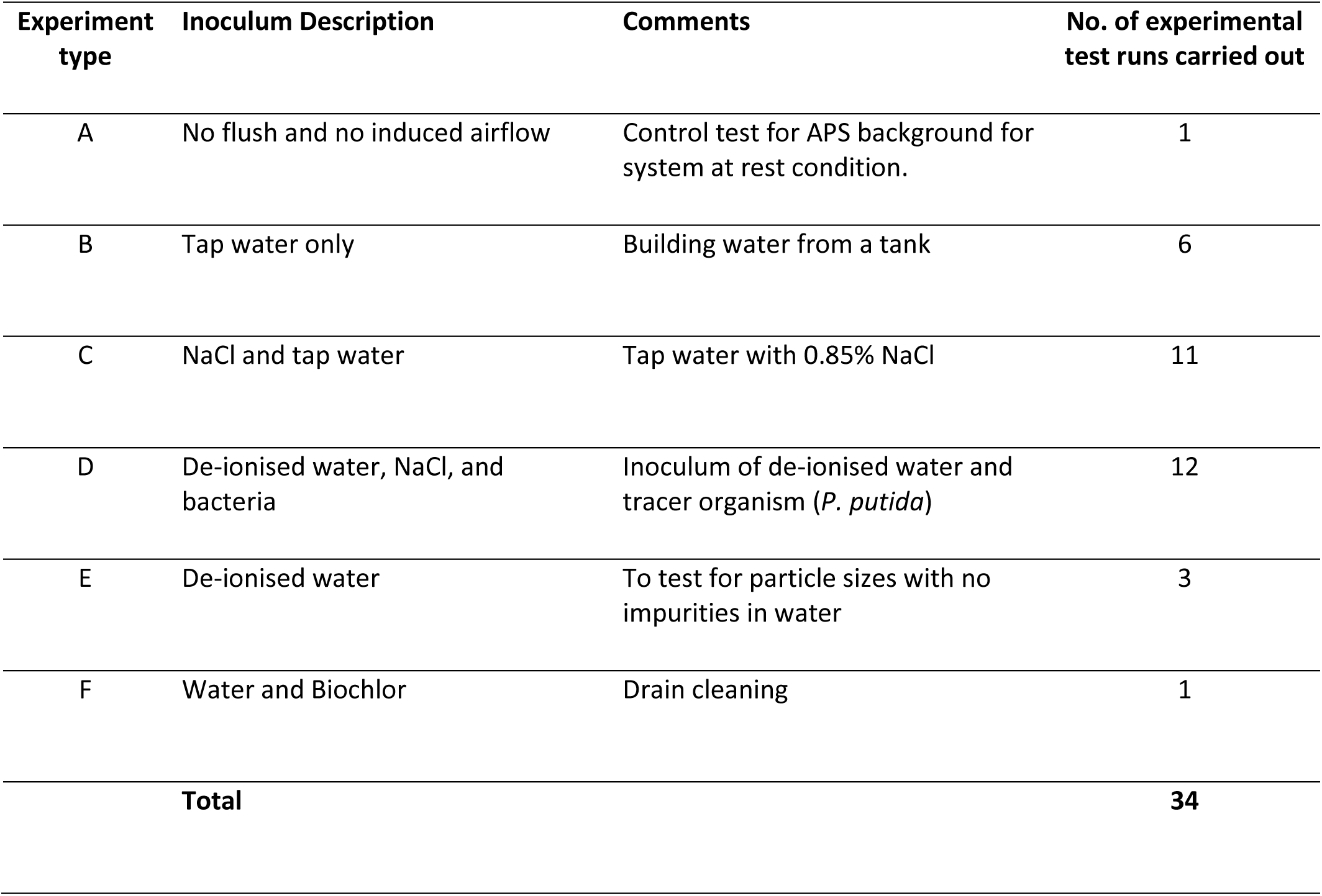
Experiments carried out.

## 3. Results

### 3.1 Aerosols / particles

APS results were compiled for all the tests carried out and analysed for: (i) total concentration of all particles (number of particles/cm^3^); (ii) total number of particles per second; (iii) distribution of particle sizes in the air samples taken at each sampler location; (iv) variation of particle size distribution at the different sampler locations; and (v) effect of flush water volume on particle size distribution.

#### 3.1.1 Single Flush

An example of the total number of particles per second emitted, and their concentration, for a typical test is shown in Figure 5. The figure shows the results of Experiment D, where the toilet flush liquid used was de-ionised water, NaCl, and *P. putida* bacterium (as detailed on Table 2 above), measured at APS sampler location 4 (see Figure 3). The background level is stable at just under 4 particles per cm^3^, which is expected in this type of test environment – a large water engineering laboratory. The peak particle counts resulting from the aerosolization from the flush can clearly be seen.

**Figure 5:**
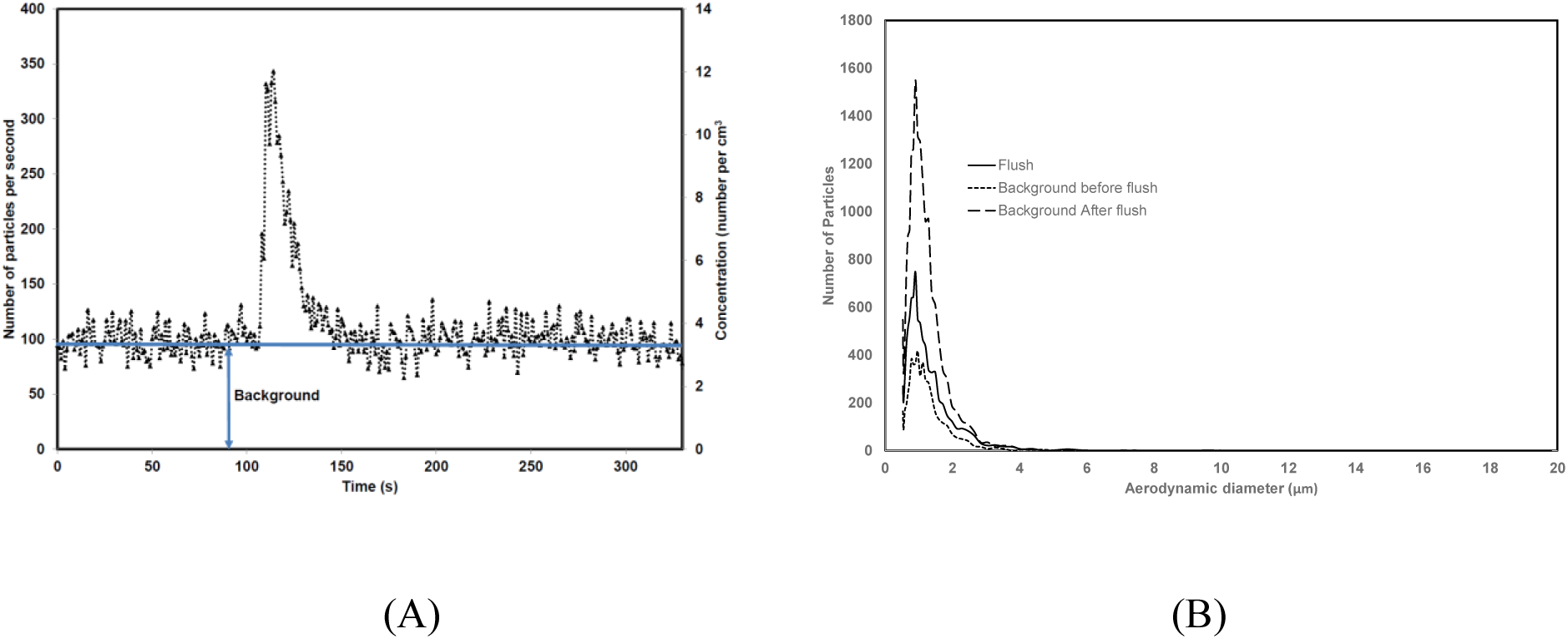
A single flush of de-ionised water, NaCl and bacteria. The background was sampled for 105 seconds before the flush was initiated and stabilised at an average 3.65 particles/ cm^3^. (A) shows particle numbers and concentration over time and (B) shows the distribution of particles sizes.

The peak of the induced spike was 245 particles/second at a concentration of 9 particles/cm^3^ (normalised without background). An average background particle level of 87 particles/second was also observed. Analysis of particle sizes show that 99.5% of all particles have an aerodynamic diameter ≤5 μm.

### 3.1.2 Background analysis (multiple consecutive flushes)

Figure 5 illustrates that there was a considerable number of particles in the background before the flush was initiated and an even greater number ≤5 μm after the flush event. While this is most likely due to the nature of the environment and operation of fans, movement of people may also have caused increased activity. In order to test the continued generation of background particles in the air, a three flush experiment was initiated using deionised water to reduce impurities in the system (one of the Experiment E tests as detailed in Table 2 above, and measured at APS sampler location 4 as detailed in Figure 3 above). The result is shown in Figure 6 below.

**Figure 6:**
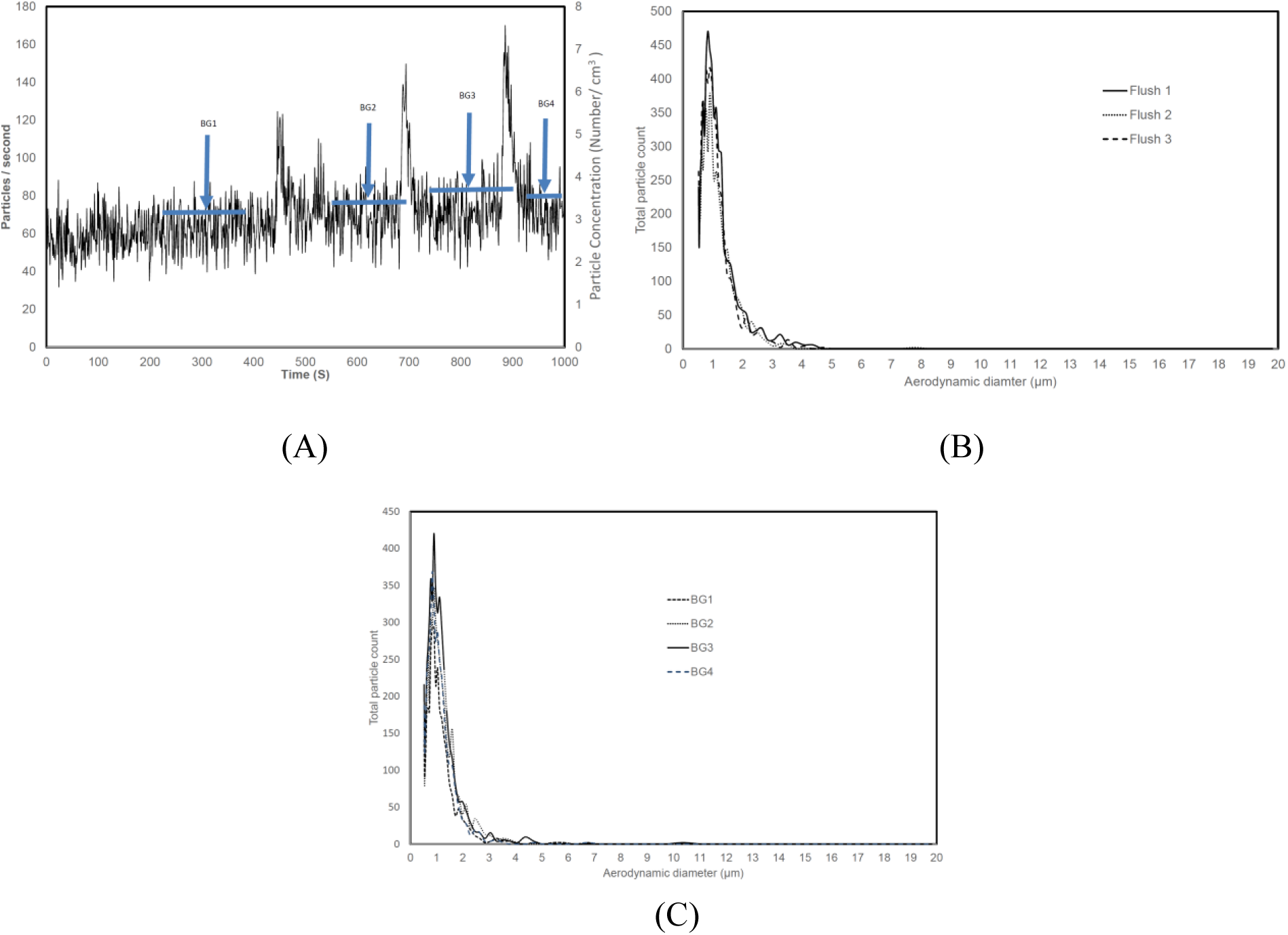
(A) Particle count and concentration for 3 consecutive flushes (de-ionised water) (B)size distribution for the particles emitted during the flush and (C) shows the size distribution for the background particles before and between flushes. (all background samples were for 53 seconds)

#### 3.1.3 Location of sampling

To establish if the characteristics of the particle emissions (particularly size distribution) differed throughout the system, a series of experiments were carried out sampling at all four locations (APS sampler locations 1-4 as detailed in Figure 3 above). The results of the size distributions are shown in Figure 7 below.

**Figure 7:**
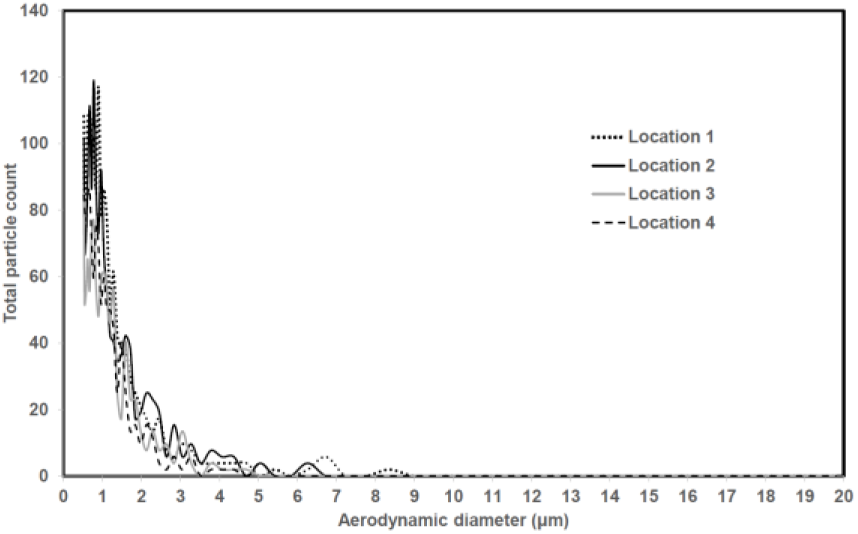
Particle size distribution at APS sampler locations 1-4 (see Figure 3). APS sampler location 1 is closest to the toilet flush and APS sampler location 4 furthest from the toilet flush.

The particle size distribution at each of the four APS sampler locations can be seen to follow the same general form of a single flush profile with the majority of particles being ≤ 5μm in size, with only a very small number of larger diameter particles (> 5μm). There were no particles with an aerodynamic diameter greater than 9 μm detected in the system following the toilet flush and none greater than 11 μm in the background data. (the particle size limit of the system being 20 μm)

#### 3.1.4 Flush Volume

Most of the tests were carried out using a simulated toilet flush volume of 6 litres. An additional set of experiments were carried out using a reduced volume to see the impact on aerosol generation, Figure 8 below shows the results.

**Figure 8:**
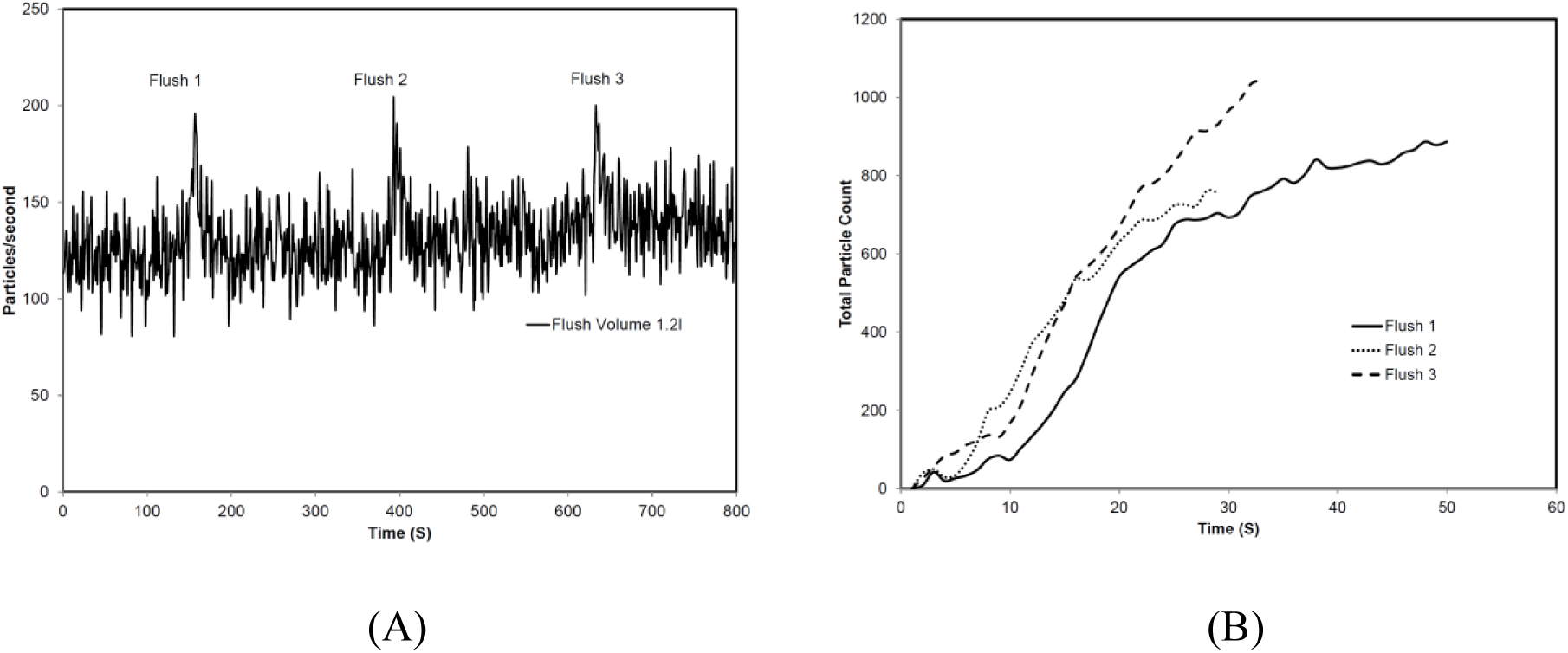
Particle results for reduced flush volume (1.2l) The spikes associated with each flush are shown in (A) with an average normalised peak emissions of 73 particles/second. Cumulative emissions for the three flushes are shown in (B) with cumulative emissions between 758 and 1045 particles.

### 3.2 Viable bioaerosol dynamics

An example slit-to-agar PIA plate after incubation is shown in Figure 9. The colonies start appearing a short time after the flush is initiated. This time is determined by the delay between turning on the slit-to-agar sampler and the initiation of the toilet flush and the time taken for the tracer organism to travel from the discharge point to the slit-to-agar sampler, approximately 10 metres. The pattern of colonies shows clustering at the beginning, shortly after the flush is initiated with a tailing off until the event ceases to contribute further tracer organism. An analysis of the kinetics of viable quantities of the tracer organism is given below.

**Figure 9:**
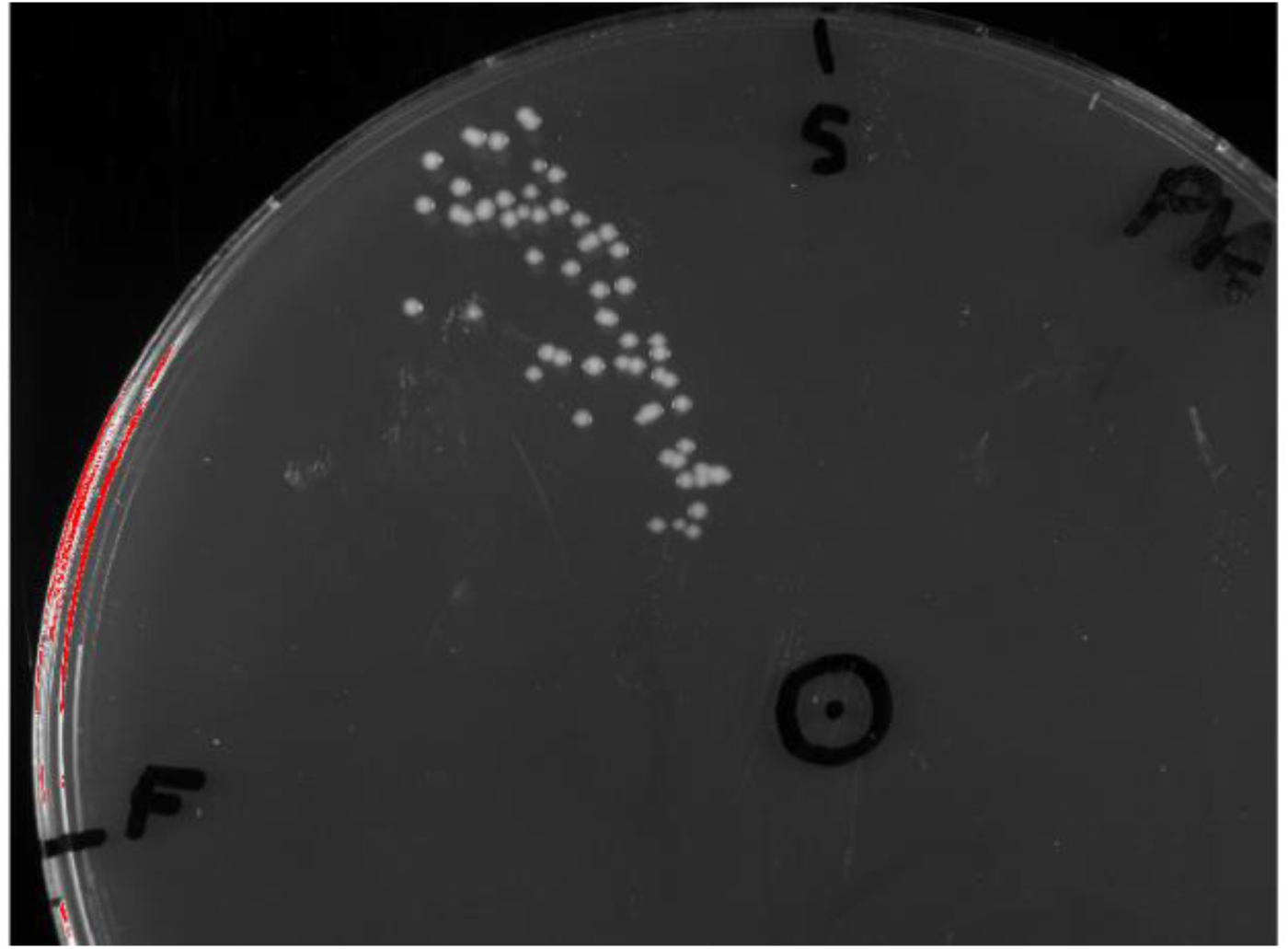
Image of 150 mm diameter PIA plate taken from the slit-to-agar sampler (Experiment type D). Start (S) and finish (F) of 5 min sampling indicated.

Analysis of one slit-to-agar plate is presented in Figure 10. The toilet flush, instigated at 60 seconds, lasted for approximately 10 seconds (see Figure 4 above). The highest numbers of CFUs can be seen to exit the ducting between 75 and 90 seconds (15-30 seconds after the toilet flush). The number of CFUs then tail off and stop around 140 seconds (80 seconds after the flush) A summary of all the experiments carried out is shown in Table 3. In addition to the CFU data, an estimate of the velocity of the tracer organism in the system is given. This is based on the time from the flush peak to the time the peak CFUs occurred. It can be seen that the tracer organism velocity is much lower than the air velocity.

**Table 3:** Summary of experimental results on bacteria transport mechanisms.

| Experiment No. | Total CFU | Peak CFU | Time of Peak count (secs) | Total Duration (secs) | Air velocity (m/s) | Estimated* bacteria velocity (m/s) |
| --- | --- | --- | --- | --- | --- | --- |
| 1 | 79 | 21 | 3.4 | 155 | 6.03 | 3.38 |
| 2 | 41 | 8 | 19.3 | 82.41 | 5.30 | 0.60 |
| 3 | 57 | 9 | 18.34 | 76.65 | 5.30 | 0.63 |
| 4 | 18 | 4 | 9.72 | 88.53 | 5.30 | 1.18 |
| 5 | 9 | 2 | 14.46 | 147.6 | 6.03 | 0.80 |
| 6 | 40 | 10 | 23.28 | 68.28 | 3.39 | 0.49 |
| 7 | 15 | 5 | 20 | 48.73 | 5.30 | 0.58 |

\* Bacteria velocity is an estimate based on the time from the flush peak to the time the peak CFUs occurred.

**Figure 10:**
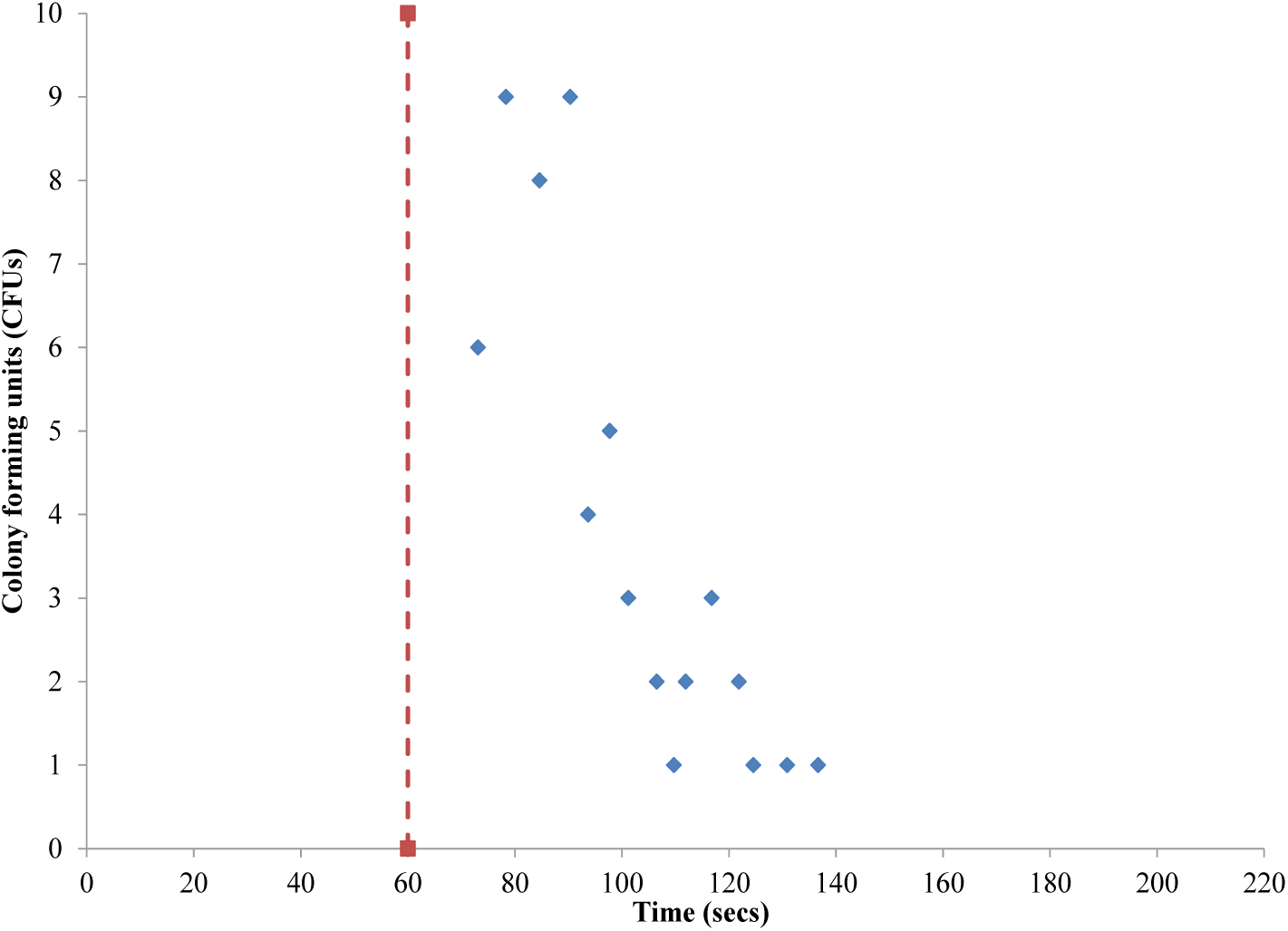
Example CFUs against time for a D type experiment.

**Figure 11:**
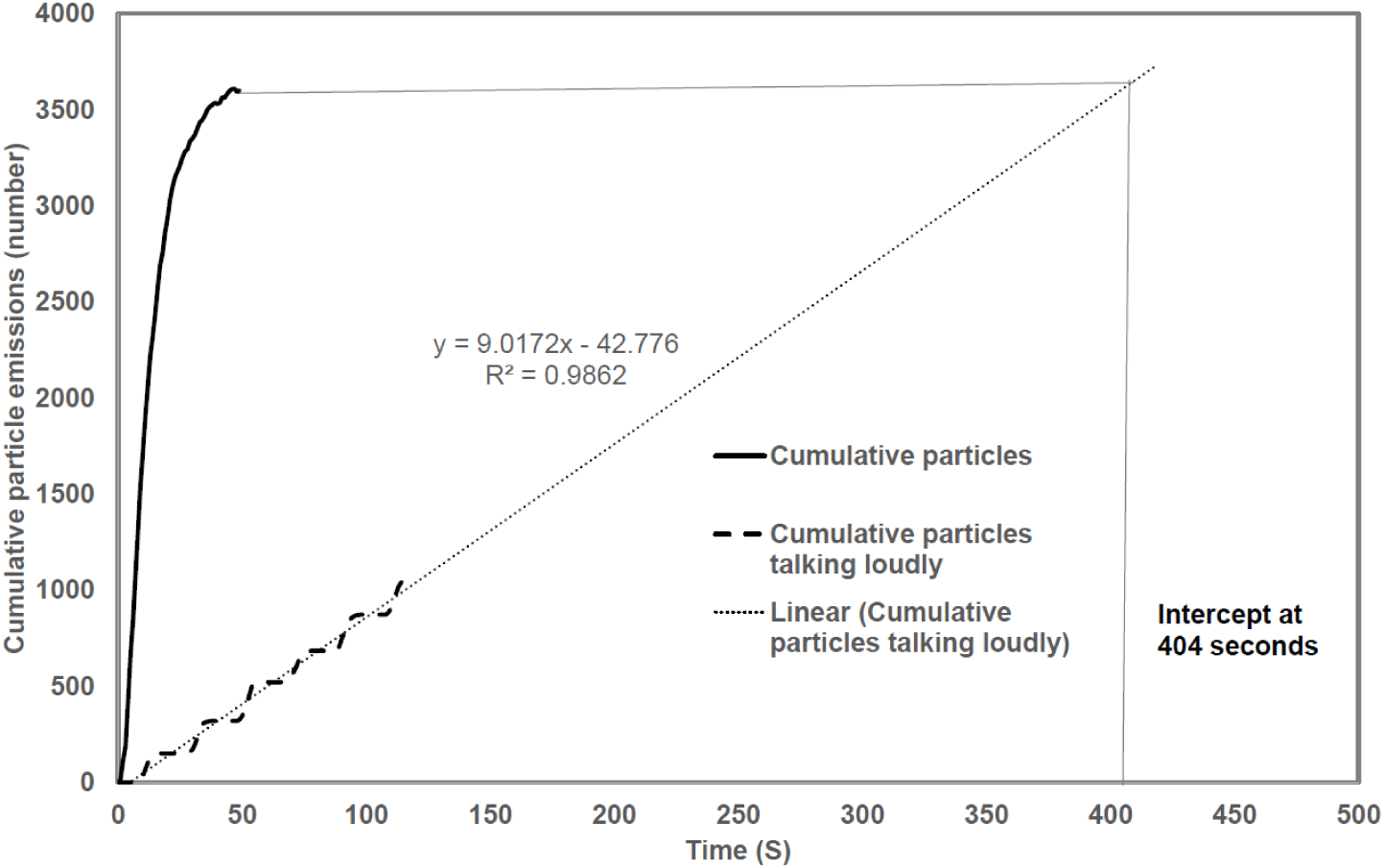
Equivalence between particles emitted as a result of a toilet flush in a sanitary plumbing system and talking loudly. Data adapted from Asadi et al ^34^.

## 4 DISCUSSION

### 4.1 General

The results of the slit-to-agar method (Experiment D) show that a tracer organism flushed into the sanitary plumbing system can be detected in the extract duct of a room on the level above that of the contaminant input level within a building. The APS data shows a profile consistent with the shape of the initiating toilet flush, but over a longer period of time. Typically, a 10 second toilet flush event was found to result in aerosols being emitted into the building over 60 seconds, however, this was found to range from 30-110 seconds in duration. This profile is repeated in the slit-to-agar results for the tracer organism, with a characteristic sharp rise in numbers followed by a longer tail off.

Although slit-to-agar samplers are typically used to quantify viable microorganisms within a given volume of air and are generally used for clean room and production area contamination monitoring ^29,30^, more recently they have found application for more general indoor air and environmental monitoring ^31,32^. Here, we show the novel application of a slit-to-agar sampler to determine the kinetics of viable cells transported in the airstream through a building’s sanitary plumbing system. The sampler effectively determined a ∼15 second time window when the highest number of cells went through the system, followed by a tail of decreasing cell numbers with time. This pattern in terms of peak and tails time was consistent across replicate experiments and with different air flow rates. The CFUs measured at the system exit ranged in duration between 48 seconds and 155 seconds.

When higher flow rates were used we found the same peak and tail patterns, however, isolated colonies also appeared 140 seconds after the toilet flush on more than one occasion. We believe this was the result of cells detaching from within the sanitary plumbing system due to shear stress under this higher air flow rate.

In our experiments, the slit-to-agar sampler was used as a semi-quantitative manner. The reason for this was that the air flow rates within the sanitary plumbing system were in excess of the sampling rate of the sampler. In addition, the duct outlet was larger than the inlet of the sampler and designed as such so as not to restrict airflow within the sanitary plumbing system. The results obtained from the slit-to-agar sampling are indicative of the spatial and temporal distribution of viable bacteria into the building from the sanitary plumbing system. While this serves as semi-quantitative data, there is evidence that slit-to-agar sampling underestimates the number of bacteria against other methods ^33^

### 4.2 APS Results in Context

In order to establish an equivalence with a known source of bioaerosol infection emission, we adapted data from Asadi *et al*., ^34^ who investigated particle emissions from people talking loudly. Asadi’s team asked subjects to talk loudly into a particle sampler to capture the magnitude, concentration and particle size distribution from a test speech involving talking loudly followed by periods of breathing only. We adapted this data to show the cumulative emissions from talking loudly for 120 seconds. This is shown in Figure 10. We also plotted the cumulative emissions from our Experiment D which used 6 litres as the toilet flush volume, and measured at APS sampler location 4. By extending the trendline on the Asadi data we can see that the number of particles emitted from the sanitary plumbing system as a result of a toilet flush is equivalent to a person talking loudly for 404 seconds (or just over 6 and a half minutes).

## 5 CONCLUSIONS

The experiments show that the generation and transport of aerosols and a tracer organism, *P. putida* KT2440, within a sanitary plumbing system are determined by the unsteady characteristics of the air and water flows within the system itself. The pattern of particles recorded by APS and the bacteria cultured on the PIA plates are consistent with the pattern of the toilet flush mechanism which initiates the inflow of the tracer organism into the system, and, sets up the turbulent conditions from which aerosolisation occurs.

The characteristics of the particles generated within the sanitary plumbing system following a simulated toilet flush are almost entirely aerosols with an aerodynamic diameter ≤5 μm. There were only a small number of particles between 10 and 11 μm when measured nearer the source. Over 99.5% of all particles were <5 μm in diameter. These characteristics mean that aerosols and bioaerosols from a sanitary plumbing system are amenable to be carried long distances on airflows inside buildings. We assume that larger droplets were generated but were not carried on the airflow and were deposited on pipe walls and in the chamber. The total number of particles emitted by the system into the room has been shown to be the equivalent of a person taking loudly for approximately 6 and a half minutes.

Particle count was influenced by flush volume, but it was not possible to determine if there was any direct influence from airflow rate, although it may have had some effect on the duration of the emission. A wider range of airflow rate would be required to verify this. Typical emissions resulting from a 6 litre flush was in the range 280 – 400 particles per second at a concentration of typically 9 to 12 number per cm^3^ and a total particle count in the region of 3,000 to 4,000 particles, whereas the peak emissions from a 1.2 litre flush was 60 - 80 particles per second at a concentration of 2.4 to 3 number per cm^3^ and a total particle count in the region of 886 to 1045 particles. The reduction in particles is in direct proportion to the reduction in flush volume.

The event-causing bioaerosols in these experiments was a simulated toilet flush containing the *P. putida* KT2440 tracer organism at a dose rate of ∼10^6^ cfu ml^-1^. This event lasted no more than 10 seconds. The discharge of the toilet flush into the sanitary plumbing system instigated an aerosolisation process due to the turbulent flow conditions. The resulting CFUs measured at the system exit ranged in duration between 48 seconds and 155 seconds. The pattern of CFU on the slit-to-agar sampler PIA plates confirms that the tracer organism concentration followed the flush characteristic (in litres/sec), the elongation of the surge wave being attributable to the difference in velocity between the bioaerosols and the air.

This work highlights the complexity of tracking and quantifying aerosols and bioaerosol transmission due to short-burst events such as those found in sanitary plumbing systems. The current COVID-19 pandemic has emphasised the challenges for all building systems in maintaining healthy indoor environments and highlights the consequences of ignoring potential disease transmission routes.

## Data Availability

Data is available upon request from the corresponding author.

## ACKNOWLEDGEMENT

This research was funded by the School of Energy, Geoscience, Infrastructure and Society research seed-corn fund at Heriot-Watt University

## 8 AUTHOR CONTRIBUTIONS

All authors contributed equally to the design, collection and analysis of the data. MG secured the funding and all authors contributed equally to the preparation of the manuscript.

